# Real-World Effectiveness of Nirsevimab Against Respiratory Syncytial Virus: A Test-Negative Case-Control Study

**DOI:** 10.1101/2024.09.12.24313545

**Authors:** Hanmeng Xu, Camilla Aparicio, Aanchal Wats, Barbara L. Araujo, Virginia E. Pitzer, Joshua L. Warren, Eugene D. Shapiro, Linda M. Niccolai, Daniel M. Weinberger, Carlos R. Oliveira

**Author notes:** Corresponding author: Carlos R. Oliveira, Address: 464 Congress Avenue, Suite 204, New Haven, CT 06520. contributed equally as co-senior authors (Daniel M. Weinberger, Carlos R. Oliveira).

## Abstract

**IMPORTANCE:** Nirsevimab, a long-acting monoclonal antibody, has demonstrated efficacy against RSV-related lower respiratory tract infections (LRTIs) in clinical trials. Post-licensure monitoring is essential to confirm these benefits in real-world settings.

**OBJECTIVE:** To evaluate the real-world effectiveness of nirsevimab against medically attended RSV infections in infants and to assess how effectiveness varies by disease severity, dosage, and time since immunization.

**DESIGN, SETTING, AND PARTICIPANTS:** This test-negative case-control study used inpatient, outpatient, and emergency room data from the Yale New Haven Health System. Nirsevimab-eligible infants who were tested for RSV using polymerase chain reaction between October 1, 2023 and May 9, 2024 were included. Cases were infants with confirmed RSV infections; controls were those who tested negative.

**EXPOSURE:** Nirsevimab immunization, verified through state immunization registries.

**MAIN OUTCOMES AND MEASURES:** Effectiveness was estimated using multivariable logistic regression, adjusting for age, calendar month, and individual risk factors. Separate models examined effectiveness by clinical setting, disease severity, dose, and time since immunization. Broader outcomes, including all-cause LRTI and LRTI-related hospitalization, were also analyzed, with stratification by early and late respiratory seasons.

**RESULTS:** The analytic sample included 3,090 infants (median age 6.7 months, IQR 3.6-9.7), with 680 (22.0%) RSV-positive and 2,410 (78.0%) RSV-negative. 21 (3.1%) RSV-positive and 309 (12.8%) RSV-negative infants received nirsevimab. Effectiveness against RSV infection was 68.4% (95% CI, 50.3%-80.8%). Effectiveness was 61.6% (95% CI, 35.6%-78.6%) for outpatient visits and 80.5% (95% CI, 52.0%-93.5%) for hospitalizations. The highest effectiveness, 84.6% (95% CI, 58.7%-95.6%), was observed against severe RSV outcomes requiring ICU admission or high-flow oxygen. Although effectiveness against RSV infections declined over time, it remained significant at 55% (95% credible interval, 16%-75%) at 14 weeks post-immunization. Protective effectiveness was also observed against all-cause LRTI and LRTI-related hospitalizations during peak RSV season (49.4% [95% CI, 10.7%-72.9%] and 79.1% [95% CI, 27.6%-94.9%], respectively). However, from February to May, when RSV positivity was low, effectiveness against these broader outcomes was negligible.

**CONCLUSIONS AND RELEVANCE:** Nirsevimab provided substantial protection against RSV-related outcomes for at least three months. These findings support the continued use of nirsevimab and provide evidence that may help build public confidence in the immunization program.

**Key Points:** *Question:* What is the effectiveness of nirsevimab against medically attended respiratory syncytial virus (RSV) infections in infants?

*Findings:* 680 RSV test-positive cases and 2,410 RSV test-negative controls were included in this test-negative case-control study. Nirsevimab’s effectiveness was 69% against RSV infections, 81% against RSV-associated hospitalization, and 85% against severe RSV disease. Effectiveness against RSV infection declined from 79% at 2 weeks post-immunization to 55% at 14 weeks post-immunization.

*Meaning:* Nirsevimab provides strong protection against a wide range of RSV outcomes, but its effectiveness diminishes over time. These data can be utilized to optimize nirsevimab’s implementation and sustain its uptake.

## Introduction

Respiratory syncytial virus (RSV) is a major cause of acute lower respiratory tract infection (LRTI), particularly affecting newborns and infants under 1 year of age. Globally, RSV is responsible for approximately 1.4 million hospital admissions and 13,300 in-hospital deaths annually among infants aged 0-6 months [1]. The recent introduction of prophylactic interventions, such as nirsevimab, provides a promising strategy to mitigate RSV’s impact on this vulnerable population.

Nirsevimab, a long-acting monoclonal antibody, was licensed by the United States (US) Food and Drug Administration in July 2023 after demonstrating safety and efficacy in prelicensure trials[2]. These trials reported 79% efficacy (95% confidence interval [CI], 69–86) against medically attended RSV, 81% (95% CI, 62–90) against RSV requiring hospitalization, and 90% (95% CI, 16–99) against severe RSV requiring intensive care unit (ICU) admission[3]. Following its licensure, the Centers for Disease Control and Prevention’s (CDC) Advisory Committee on Immunization Practices (ACIP) recommended nirsevimab for infants under 8 months entering their first RSV season and for high-risk infants aged 8-19 months[4].

While pre-licensure clinical trials demonstrated strong efficacy, it is essential to validate these findings through post-licensure studies that assess nirsevimab’s effectiveness in real-world settings. Such studies are needed to ensure that the protective effects of immunizations remain as they are being used in routine clinical practice, where factors like comorbidities, access to care, and clinician practices can influence outcomes. Early real-world data from the 2023-2024 RSV season in Europe and the US show effectiveness ranging from 70% to 90% against hospitalization for RSV-associated LRTI[5–9]. However, several gaps in knowledge remain. Specifically, there are limited data on nirsevimab’s long-term effectiveness, its impact at different dosages, and its ability to prevent milder RSV cases. Furthermore, there is a need for further exploration of its effectiveness in diverse populations, particularly those with underlying health conditions. To address these gaps, this study aims to evaluate nirsevimab’s real-world effectiveness in a diverse US patient population and examine how protection varies over time, by disease severity, and by dosage used.

## Methods

### Study Design and Study Population

The effectiveness of nirsevimab against medically attended RSV infection was estimated using the test-negative case-control study design. The study population included all patients who were born after October 1st, 2022, were tested for RSV due to a suspected acute respiratory infection (ARI)[10], and received care in a facility affiliated with the Yale New Haven Health System (YNHHS) between October 1^st^ 2023 and May 9^th^ 2024. The YNHHS is the largest health system in Connecticut, and consists of five integrated hospital networks, 30 emergency or urgent care centers, and over 130 outpatient clinics from Westchester County, New York, to Rhode Island, all integrated using a single electronic health record (EHR) system.

Patients were excluded from these analyses if they were not age-eligible for nirsevimab when it became available on October 1^st^ 2023[11], or if they resided outside Connecticut, New York, or Rhode Island. The geographic restriction was implemented to ensure that immunization records could be verified through state immunization registries, which are directly integrated with the YNHHS EHR. Infants were considered eligible for nirsevimab if they were born during the season (after October 1st, 2023), or if they were under 8 months old and entering their first RSV season, or if they were between 8 and 12 months old with a risk factor for severe RSV (see Figure 1 for the detailed inclusion process and Table S1 for the definitions for risk factors).

**Figure 1.**
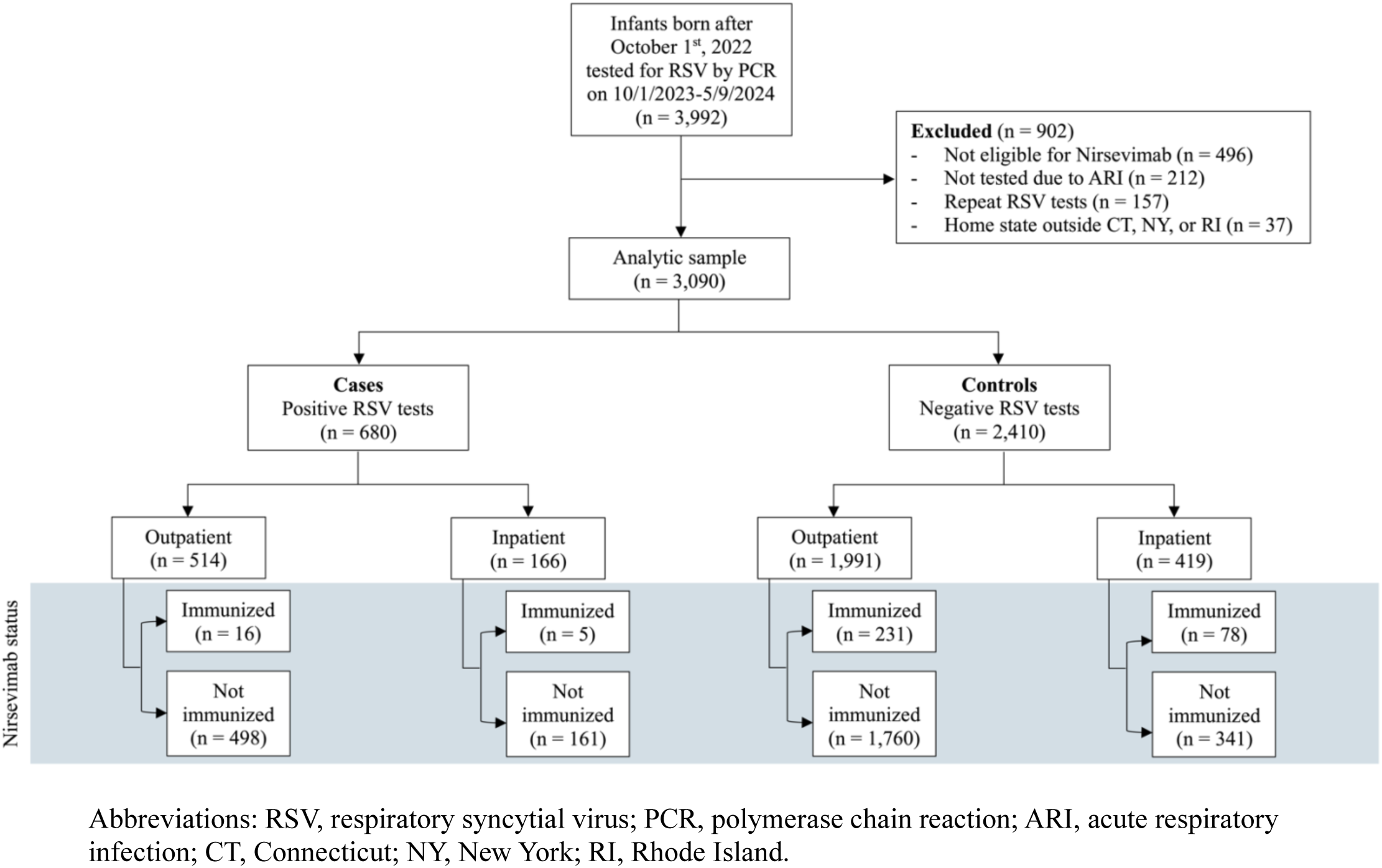
Selection of RSV test records.

### Data Sources and Study Definitions

For all the infants who met the eligibility criteria, reviews of medical records and state immunization registry searches were conducted to capture information on patient characteristics, immunization history, and potential confounders. Relevant clinical and laboratory data associated with each patient’s RSV test, such as chief complaints, problem lists, encounter diagnoses, and presence of any other acute or chronic diseases were abstracted by trained investigators from the EHR. The clinical outcomes following hospitalization were also recorded, such as hospital and ICU length of stay, and maximum respiratory support needed during hospitalization. Patient characteristics including age, race and ethnicity, gestational age, birth weight, and type of insurance were also abstracted.

Cases were defined as infants with a medically attended RSV infection confirmed by nasopharyngeal polymerase chain reaction (PCR). Controls were infants with ARI who tested negative for RSV. For a given patient, if there was more than one positive test during the study period, only the first was included in the study. If an eligible control had more than one negative test within 14 days, the first negative test was selected for this period. The primary exposure of interest in our study was the nirsevimab immunization status. Only documented immunization dates were included in the analysis. Infants were classified as “immunized” if they received a dose of nirsevimab prior to their RSV test.

### Statistical analysis

The characteristics of the study population were summarized using frequency distributions and measures of central tendency. Univariate analyses were performed to compare RSV-positive patients with negative controls, and unimmunized with immunized infants. Covariate balance between groups was assessed to detect potential confounders using standardized mean differences (SMD), with absolute SMDs of less than 0.20 indicating adequate balance.

For our primary analysis, the effectiveness of nirsevimab against medically attended RSV infection was quantified using all eligible patients in our study population. Effectiveness was calculated as one minus the odds ratio (OR) of immunization with nirsevimab among cases and controls, using logistic regression. Adjusted effectiveness estimates were derived using multivariable models that controlled for age, calendar month, and the presence of at least one risk factor. Non-collinear potential confounders were selected for the final models through backward selection based on the Akaike information criterion (AIC). Model formulas and AIC values are provided in the supplementary materials (Figure S1, Table S2).

For our secondary aims, separate models were fitted to analyze the data based on clinical setting (inpatient vs outpatient), disease severity, nirsevimab dosage used, and time from immunization. For the severity analysis, patients were considered to have severe disease if they were hospitalized within 14 days of RSV testing and required either transfer to a pediatric ICU or high levels of respiratory support during hospitalization, such as high-flow nasal cannula (≥2 liters per minute), continuous or bilevel positive airway pressure, or invasive mechanical ventilation. The extent to which the effectiveness of nirsevimab decreased over time was quantified using logistic regression within a Bayesian framework. The parameter representing the effectiveness of immunization was time-varying at bi-weekly intervals of time since immunization. These models used weakly-informative prior distributions and imposed a monotonic structure on the regression coefficients that represent nirsevimab’s effectiveness for increasing time since immunization. Medians and 95% credible intervals were calculated from the collected posterior samples, and convergence was evaluated using trace plots (Figure S6). A comprehensive description of the Bayesian models is provided in the supplementary material.

The effectiveness of nirsevimab was also assessed using broader endpoints, including all-cause lower respiratory tract infection (LRTI) and all-cause LRTI hospitalization across the entire respiratory season, with additional stratification by early (October to January) and late (February to May) periods.

### Sensitivity analyses

Several sensitivity analyses were conducted to assess the robustness of our findings. First, we assessed for differences in effectiveness when employing different exposure and outcome definitions. Specifically, we explored restricting our analysis to only medical visits where encounter diagnoses indicating lower respiratory tract infection (LRTI) were recorded as either a primary or secondary diagnosis (Table S1). We also explored restricting controls to only those who tested positive for other respiratory viruses (i.e., influenza, adenovirus, rhinovirus, parainfluenza). In terms of exposure, we explored defining patients as “immunized” if they received nirsevimab ≥7 days prior to RSV testing, as was done in earlier reports to account for the mean RSV incubation period and the time required to reach peak antibody concentration[9]. Second, we assessed whether excluding infants whose mothers received the maternal RSV vaccine or those who were born during the previous RSV season would significantly alter the results. Last, we repeated our analysis using the hepatitis B vaccine as a “sham” exposure, as previously described [12, 13]. Since the hepatitis B vaccine is recommended to be given to all newborns but does not affect the risk of RSV infection, we expect that in the absence of bias, the proportions of cases and of controls who were immunized with the hepatitis B vaccine will not be significantly different.

Further details on study definitions and statistical analysis are provided in the Supplemental Methods. All analyses were conducted in R, version 4.3.1[14]. The code for the analysis can be found at https://github.com/Hanmeng-Xu/RSV_mAb_VE.

The institutional review board (IRB) at the Yale University School of Medicine approved the study (HIC:2000036550). The funders of this study had no role in study design; collection, analysis, and interpretation of data; writing the report; and the decision to submit the report for publication.

## Results

### Study population

Between October 1, 2023, and May 9, 2024, a total of 3,992 RSV tests were performed within the YNHH system on infants born after October 1, 2023. Of these, 3,090 met our eligibility criteria and were included in the analysis (Figure 1). The analytic sample consisted of 680 RSV-positive and 2410 RSV-negative patients. Most RSV tests occurred during emergency department or urgent care clinic visits (n= 2,505, 81.1%) in the months of December and January (Figure S2). The median age at the time of RSV testing was 6.7 months (IQR 3.6 to 9.7 months). RSV-positive cases were slightly younger than RSV-negative controls (6.1 vs. 6.9 months, p = 0.007) and had a lower proportion of prematurity (11.2% vs. 14.2%, p = 0.045). However, other demographic and clinical factors, including sex, race/ethnicity, insurance type, and prevalence of comorbidities, were comparable between the two groups (Table 1).

**Table 1.** Characteristics of included cases and controls, October 1^st^ 2023 – May 9^th^ 2024.

|  | Overall, N = 3,090<sup>1</sup> | Cases, N = 680<sup>2</sup> | Controls, N = 2,410<sup>2</sup> | Standardized Mean Difference |
| --- | --- | --- | --- | --- |
| **Sex** |  |  |  | 0.05 |
| Female | 1,317 (42.6%) | 279 (41.0%) | 1,038 (43.1%) |  |
| Male | 1,772 (57.3%) | 401 (59.0%) | 1,371 (56.9%) |  |
| (Missing) | 1 (0.0%) | 0 (0.0%) | 1 (0.0%) |  |
| **Age at testing (months)** |  |  |  | -0.14 |
| Median (IQR) | 6.7 (3.6, 9.7) | 6.1 (3.4, 9.2) | 6.9 (3.7, 9.9) |  |
| **Race and ethnicity** |  |  |  | 0.13 |
| Hispanic | 1,328 (43.0%) | 280 (41.2%) | 1,048 (43.5%) |  |
| White non-Hispanic | 820 (26.5%) | 201 (29.6%) | 619 (25.7%) |  |
| Black non-Hispanic | 533 (17.2%) | 112 (16.5%) | 421 (17.5%) |  |
| Other non-Hispanic<sup>2</sup> | 161 (5.2%) | 26 (3.8%) | 135 (5.6%) |  |
| Unknown | 248 (8.0%) | 61 (9.0%) | 187 (7.8%) |  |
| **Birth weight** |  |  |  | 0.10 |
| Median (IQR) | 3,214.3 (2,824.9, 3,563.8) | 3,265.0 (2,875.0, 3,576.2) | 3,194.5 (2,805.1, 3,553.6) |  |
| N missing (% missing) | 783 (25.3%) | 184 (27.1%) | 599 (24.9%) |  |
| **Prematurity (< 37 weeks)** | 418 (13.5%) | 76 (11.2%) | 342 (14.2%) | 0.11 |
| **Pulmonary diseases** | 156 (5.0%) | 26 (3.8%) | 130 (5.4%) | 0.07 |
| **Cardiac diseases** | 152 (4.9%) | 30 (4.4%) | 122 (5.1%) | 0.03 |
| **Anemia** | 94 (3.0%) | 15 (2.2%) | 79 (3.3%) | 0.07 |
| **Having at least one risk factor<sup>3</sup>** | 750 (24.3%) | 150 (22.1%) | 600 (24.9%) | 0.07 |
| **Insurance type** |  |  |  | 0.09 |
| Private | 983 (31.8%) | 231 (34.0%) | 752 (31.2%) |  |
| Public | 2,088 (67.6%) | 442 (65.0%) | 1,646 (68.3%) |  |
| Uninsured | 19 (0.6%) | 7 (1.0%) | 12 (0.5%) |  |
| **mAb status** |  |  |  | 0.37 |
| No | 2,760 (89.3%) | 659 (96.9%) | 2,101 (87.2%) |  |
| Yes, 100mg dose | 95 (3.1%) | 6 (0.9%) | 89 (3.7%) |  |
| Yes, 50mg dose | 235 (7.6%) | 15 (2.2%) | 220 (9.1%) |  |

<sup>1</sup>n (%)<sup>2</sup>Including Asian, Pacific Islander, Middle Eastern or Northern American, American Indian or Native American by self-reporting.<sup>3</sup>Have at least one of the following conditions recorded in the infant's medical history or diagnosis records: 1) Anemia; 2) Immunodeficiency (e.g. transplantation history, leukemia, etc.); 3) Cardiac diseases (including congenital heart diseases diagnosed at birth or any reporting of heart conditions); 4) Pulmonary diseases; 5) Down syndrome; 6) Small for gestational age (birth weight < 2,500 grams); 7) Prematurity (gestational age less than 37 weeks).

The overall uptake of nirsevimab in the study sample was 10.7% (330/3,090). Among those who received nirsevimab before RSV testing, 71.2% (n = 235) received the 50 mg dose, while 28.8% (n = 95) received the 100 mg dose. Correlates of immunization are detailed in Table S3.

Overall, 24.4% (166/680) of RSV-positive cases resulted in hospitalization. Among those hospitalized, 58.4% (97/166) required more than two liters of respiratory support, and 13.8% (23/166) required admission to the ICU (Table S4).

### Effectiveness of nirsevimab

The adjusted effectiveness of nirsevimab against any medically attended RSV infection was 68.4% (95% CI: 50.3-80.8%). Effectiveness was 61.6% (95% CI: 35.6-78.6%) for preventing RSV-associated outpatient visits, 80.5% (95% CI: 52.0-93.5%) for preventing hospital admissions, and 84.6% (95% CI: 58.7-95.6%) for preventing severe RSV (Figure 2). These findings aligned with existing clinical trials and observational data (Figure S3). The effectiveness estimates for the 100 mg and 50 mg doses were comparable, with overlapping confidence intervals (Figure S4).

**Figure 2.**
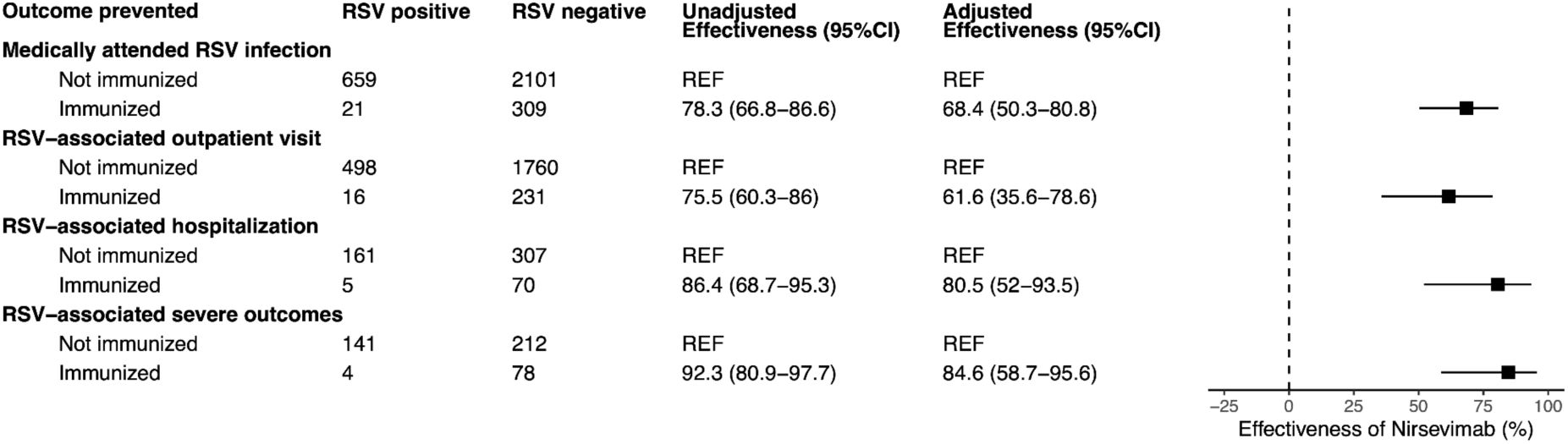
Effectiveness of nirsevimab against medically attended RSV, by clinical setting and severity. Adjusted models controlled for age, calendar month, and the presence of at least one risk factor (see Table S2).

Nirsevimab effectiveness against medically attended RSV infection decreased from 79% at 2 weeks post-immunization to 55% at 14 weeks post-immunization. A similar pattern of waning effectiveness was observed across different outcomes (Figure 3); our results were comparable to those derived from clinical trial data (Figure S5).

**Figure 3.**
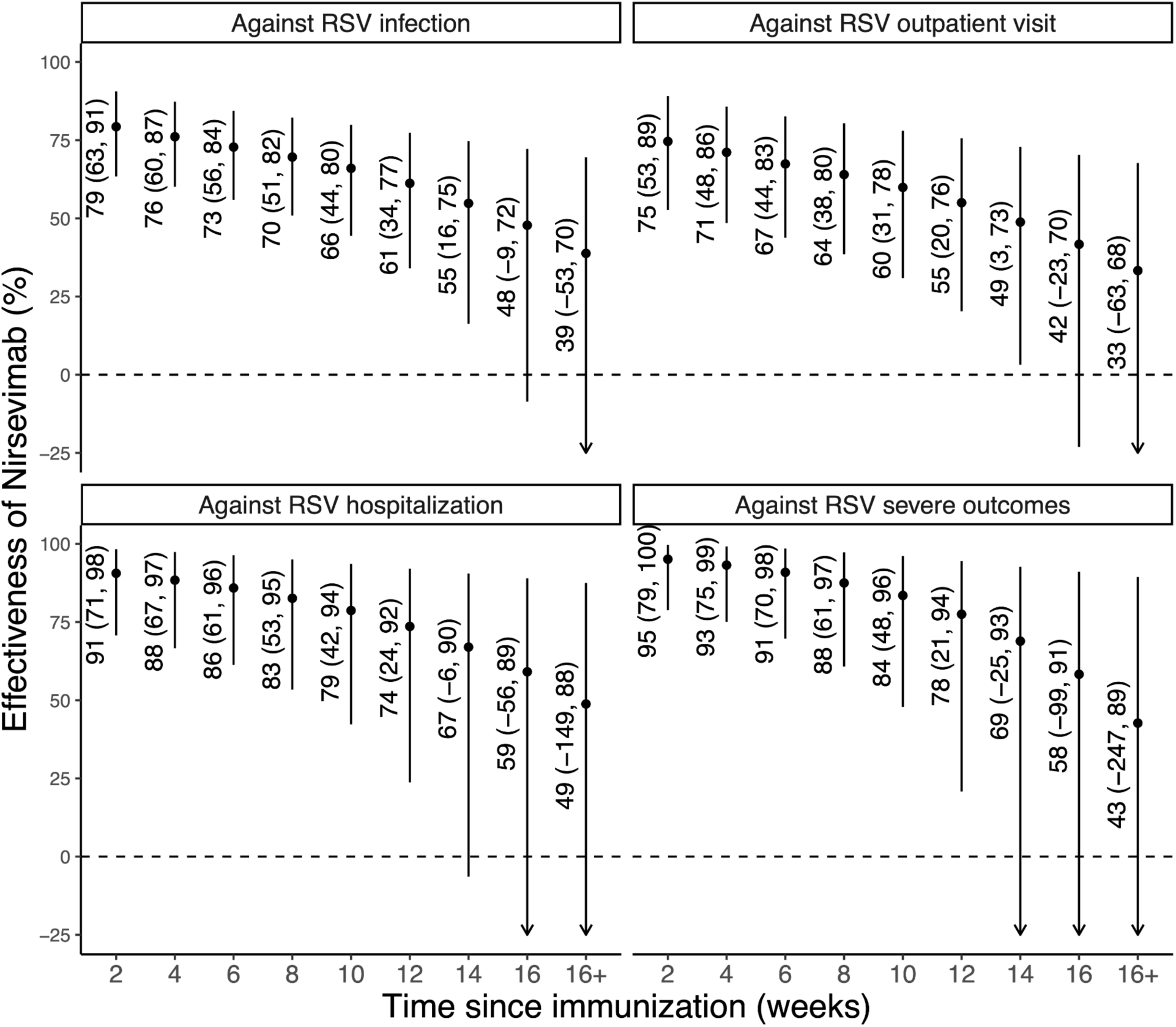
Effectiveness of nirsevimab by time since immunization. The dots represent the median estimates of effectiveness of nirsevimab in preventing the various clinical outcomes, and the error bars indicate the 95% credible intervals of the estimates.

Protective effectiveness was observed against all-cause LRTI and all-cause LRTI hospitalization during the peak months of the RSV season (49.4% [95% CI: 10.7-72.9%] and 79.1% [95% CI: 27.6-94.9%] during November and December, respectively), comparable to results from other studies (Figure 4). Given that RSV was the predominant virus during these months (RSV positivity rate 39.3%), the above estimate largely reflects the effectiveness of nirsevimab against RSV. In contrast, between February and May, when the RSV positivity dropped below 3.9%, there was no effectiveness of nirsevimab against all-cause LRTI and all-cause LRTI hospitalizations (Figure 4).

**Figure 4.**
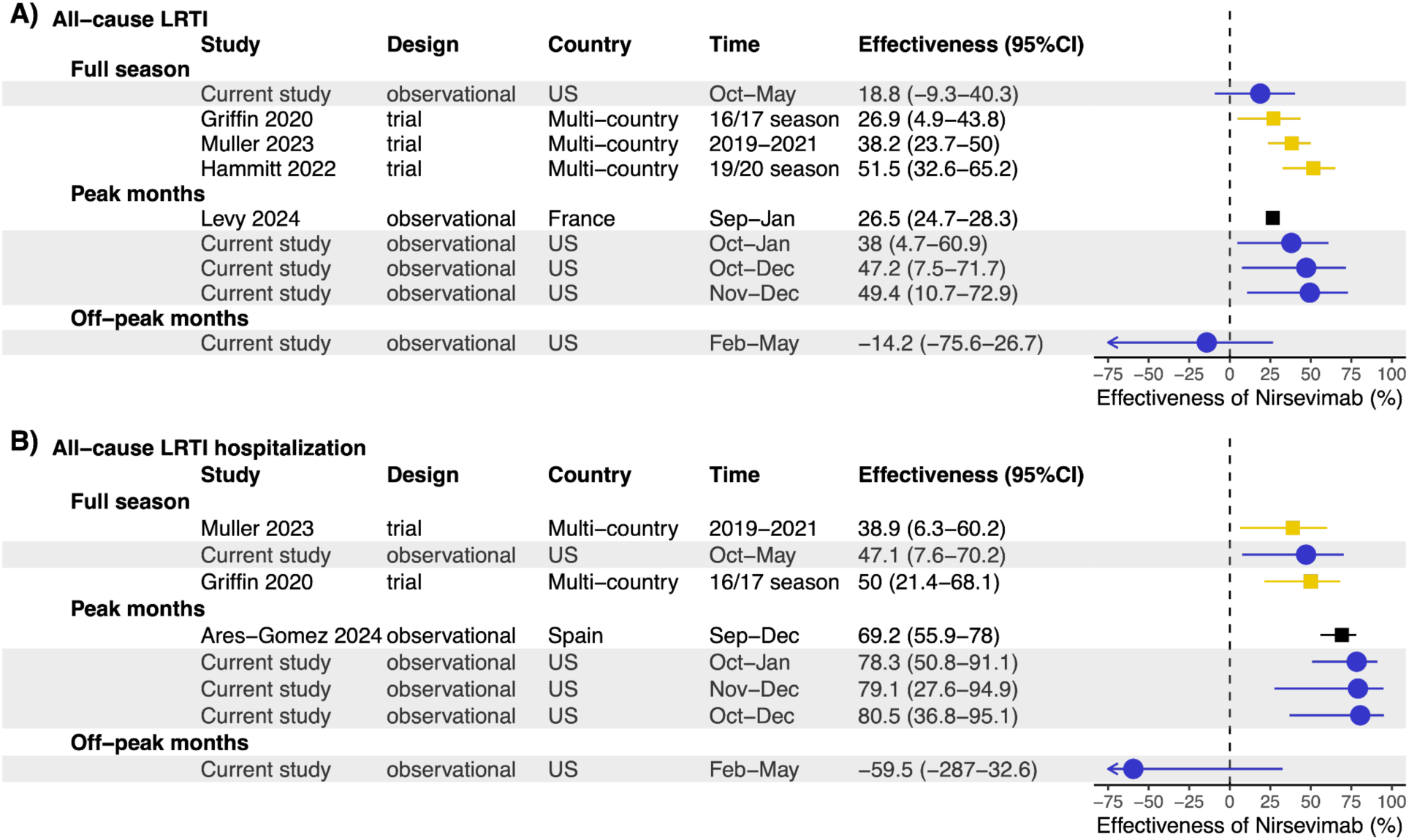
Forest plot of nirsevimab effectiveness against (A) all-cause LRTI and (B) all-cause LRTI hospitalization, stratified by time. Current study estimates are shown in blue. For comparison, estimates from three previous studies are also included. Pre-licensure clinical trial estimates are shown in gold, and other post-licensure study estimates are in black. Estimates were stratified by time (full season[30, 34, 35], peak months[5, 21], off-peak months) Only the estimate for age group 3-12 months was shown for Levy et al. 2024[21].

All sensitivity analyses generated consistent results, with less than 10% differences in point estimates of effectiveness (Table S5). As expected, the proportions of cases and controls that received the hepatitis B vaccine were nearly identical (54% and 57%, respectively, p=0.18), and the corresponding effectiveness of the hepatitis B vaccine against RSV was not statistically significant (Table S5).

## Discussion

In this study, we provide robust evidence supporting the real-world benefits of nirsevimab, with an adjusted effectiveness of 68% against medically attended RSV infections. Our data indicates that effectiveness was higher for RSV-related hospitalizations (80.5%) and severe RSV disease (84.6%). These findings align with the pre-licensure clinical trials, which reported 77-83% efficacy against RSV-related hospitalizations[15]. Emerging evidence from post-licensure studies, including a recent meta-analysis that estimated the effectiveness of 88.4% (95% CI: 84.7–91.2%) against RSV-related hospitalizations[16], further supports the findings of this study.

Our study makes several important contributions to the existing literature. First, we measure the protective effect of nirsevimab in a diverse US patient population where historically minoritized racial and ethnic groups make up the majority (>50%) of the study sample. This is notable because most previous effectiveness estimates have come from studies conducted primarily in European countries, which have distinct racial and ethnic compositions, different socioeconomic contexts, and considerably higher nirsevimab coverage. For instance, fifteen out of sixteen real-world effectiveness studies to date have been conducted in Western Europe[5–8, 17–27], where nirsevimab coverage in the target population often exceeded 70%.

Second, previous analyses, including the only US-based report [9], have primarily focused on RSV-associated hospitalizations, with limited evaluation of effectiveness against medically attended outpatient visits, which represent a substantial portion of the RSV burden [28, 29]. Our study addresses this gap but also extends the evaluation of nirsevimab to its impact on broader outcomes, such as all-cause LRTI. Prior post-licensure studies reported effectiveness of 69.2% (55.9–78.0%) against all-cause LRTI hospitalizations[5], nearly double the 39% efficacy observed in Phase III trials against these non-specific outcomes[30]. Our findings align more closely with clinical trial data, and, notably, we found no significant protective effect against all-cause LRTI outside the peak RSV season when a negligible proportion of LRTIs were due to RSV.

Third, our study provides valuable insights into the temporal dynamics of nirsevimab’s effectiveness. Given that our study spanned the entire RSV season, we were able to assess how nirsevimab’s effectiveness wanes over time—a factor less emphasized in earlier studies with shorter observation periods. While effectiveness did decline over time, the protective effects remained statistically significant for at least 14 weeks post-immunization. This waning pattern is consistent with what is known about the pharmacokinetics of monoclonal antibodies and the natural decay of passive immunity[31]. The observed decline in effectiveness beyond 14 weeks, though preliminary and based on estimates with wide uncertainty intervals, underscores the importance of timing in administering nirsevimab, especially in regions with prolonged RSV seasons.

Our study has several limitations. First, the prioritization of high-risk infants for immunization during the early roll-out phase may have introduced confounding by indication[32]. However, we adjusted for this potential effect in our analysis and conducted several sensitivity analyses that suggest residual confounding by unmeasured factors is less likely. Second, although our study benefited from a large sample size, the low uptake of nirsevimab resulted in limited statistical power and wide confidence intervals (CIs) for certain comparisons, such as effectiveness by the number of doses received. Third, because few cases were immunized more than 14 weeks before RSV testing, the effectiveness estimates beyond this period have wide credible intervals and should be interpreted with caution. Fourth, under-ascertainment of immunization or prior infections may have biased our results toward the null. However, the uptake of nirsevimab in our study is very similar to that reported by other US-based studies and CDC coverage estimates for Connecticut (7.7% uptake) [9, 33]. Furthermore, our “sham exposure” analysis suggests that our estimate of nirsevimab’s effectiveness was not likely confounded by factors associated with immunization ascertainment.

### Conclusion

This study confirms that nirsevimab is highly effective at preventing RSV-associated outpatient visits, hospitalizations, and severe disease requiring ICU admission or high-flow oxygen. Its effectiveness persisted for at least three months post-vaccination, consistent with results from randomized trials. These findings reinforce the benefits of RSV immunoprophylaxis and support US guidelines recommending nirsevimab for all infants entering their first RSV season.

## Data Availability

External researchers can make written requests to the corresponding author for sharing of completely de-identified and aggregate-level data. Data are available for researchers to allow replication of results provided all ethical and legal requirements are met. Requests will be assessed on a case-by-case basis in consultation with the lead and co-investigators. All data sharing will abide by rules and policies defined by the involved parties. Data-sharing mechanisms will ensure that the rights and privacy of individuals participating in research will be protected at all times. The study protocol and statistical code used are also available on request from the corresponding author.

## Supplementary Materials

### SECTION 1. SUPPLEMENTAL FIGURES REFERENCED IN THE MAIN MANUSCRIPT

**Figure S1.**
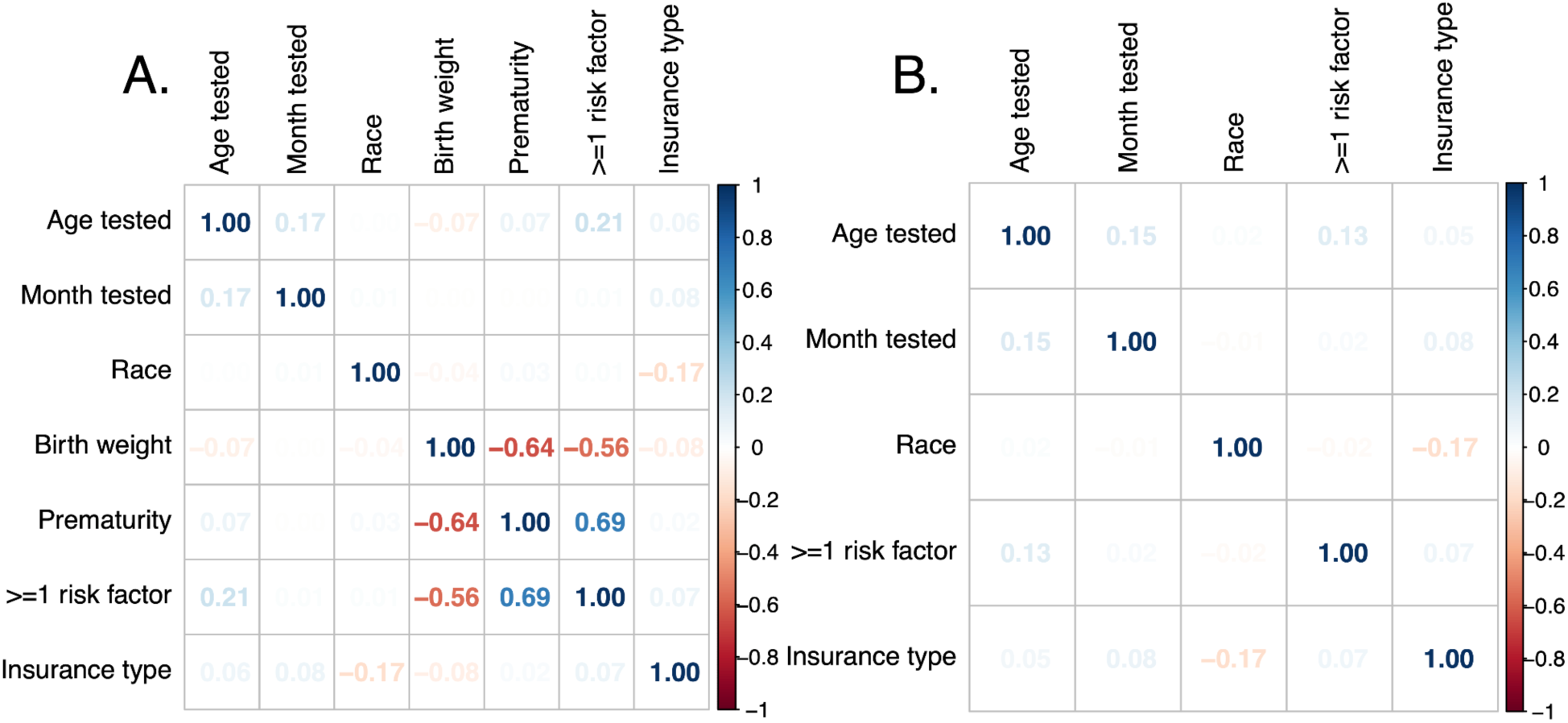
Correlation between potential confounders in the test-negative case-control analysis. Multivariate logistic regression was used to estimate nirsevimab effectiveness against various clinical outcomes. Potential confounders were selected using backward selection from variables in the initial model (panel A), including age at testing (<3, 3-5, 6-8, 9-11, ≥12 months), calendar month of testing, race/ethnicity, birth weight, prematurity (gestational age <37 weeks), presence of at least one risk factor (see Table S1), and insurance type (private, public, uninsured). The numbers show the correlation coefficients between any of the two variables, and a value larger than 0.5 was defined as a moderate or strong correlation. Due to collinearity between low birth weight and prematurity, and high rates of missing data (∼25%), only the “at least one risk factor” variable was retained (panel B).

**Figure S2.**
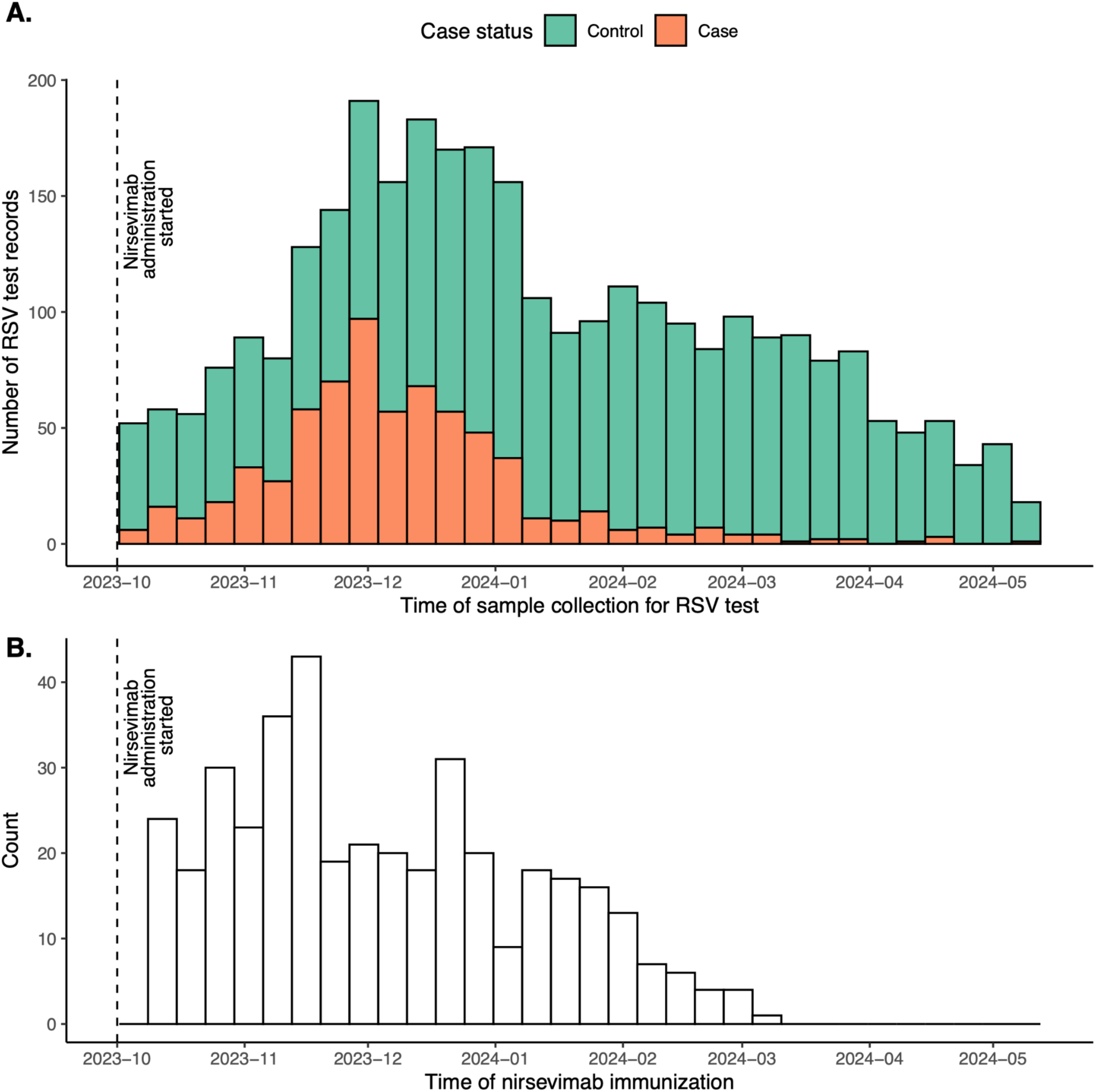
RSV tests and nirsevimab doses during the study period. Panel A shows the number of RSV tests, with orange bars for RSV-positive cases and green bars for RSV-negative controls. Panel B displays the number of nirsevimab doses administered. The vertical dashed line marks the start of nirsevimab administration in Connecticut (October 1, 2023).

**Figure S3.**
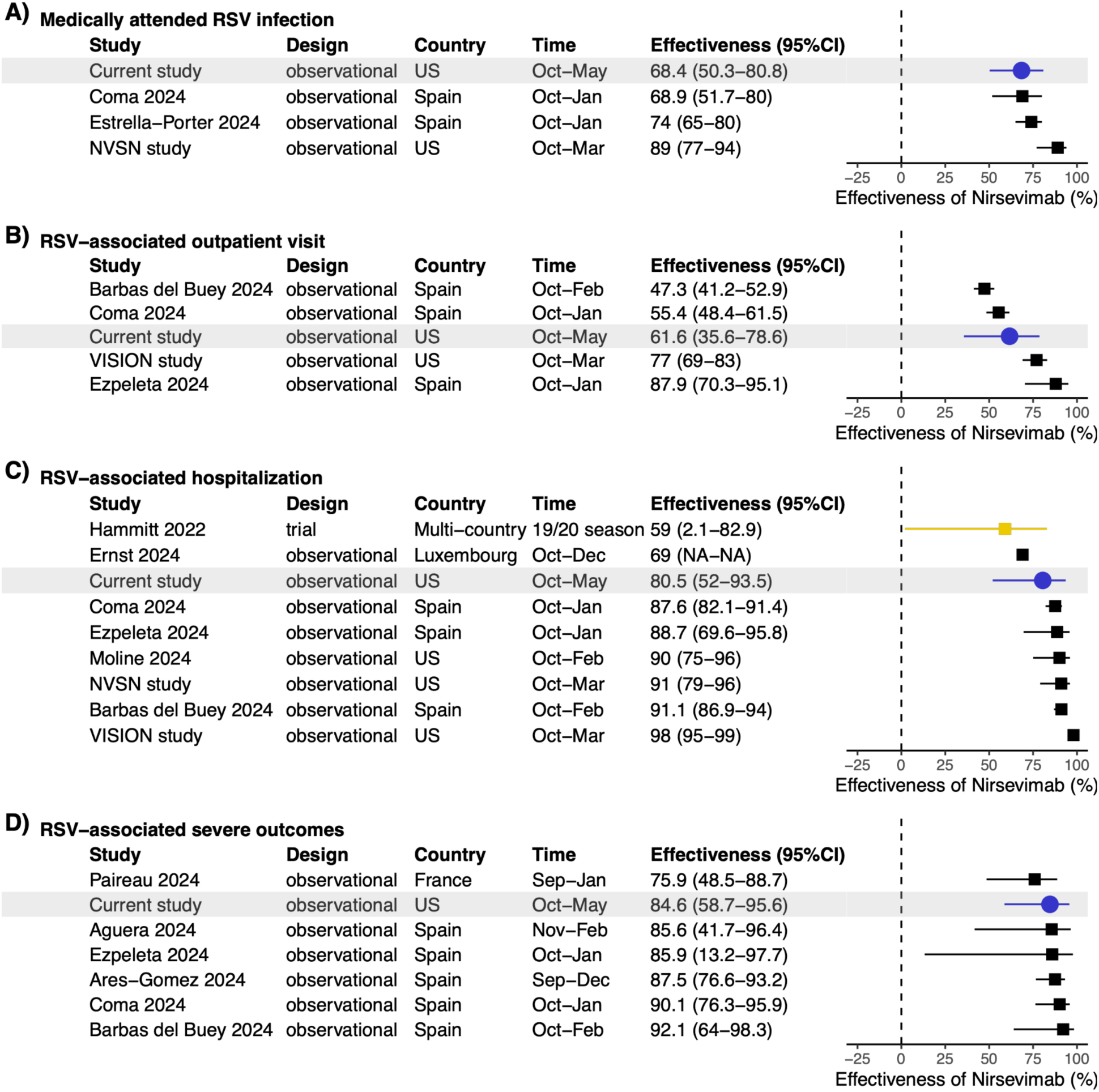
Overview of nirsevimab effectiveness: current study estimates in context with prior research. This figure contrasts adjusted effectiveness (post-licensure) and efficacy (pre-licensure) estimates from prior studies with those from the current study. The right panel shows means (dots) and uncertainty intervals (bars). Gold squares represent Phase IIb/III trial data, black squares represent observational studies, and blue circles represent the current study (highlighted in gray). Ernst et al. 2024[1] did not report uncertainty intervals.

**Figure S4.**
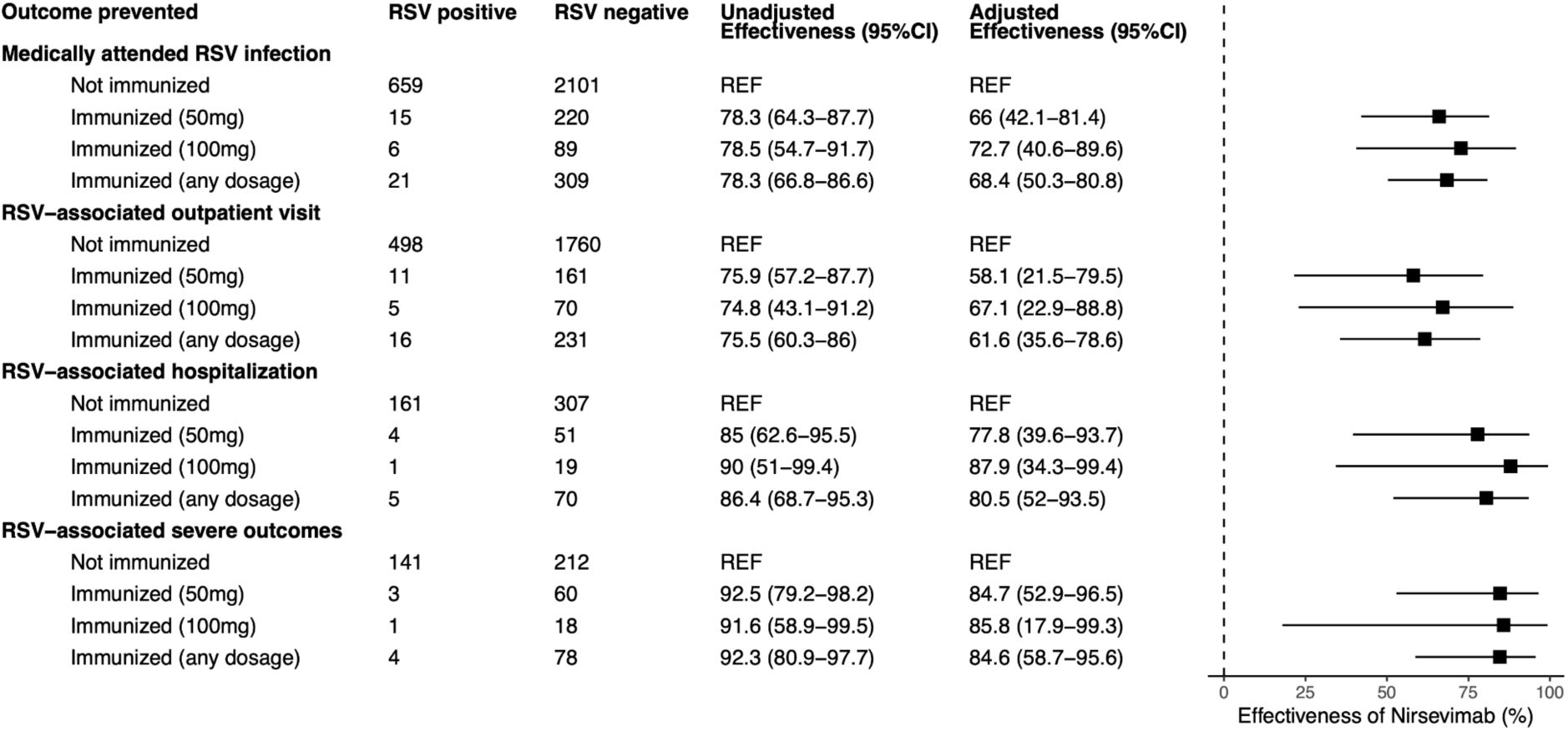
Effectiveness of nirsevimab against RSV infections by dose, clinical setting, and disease severity. Square dots indicate mean effectiveness estimates, with horizontal lines representing 95% confidence intervals. All models adjusted for age and calendar month. Models for RSV-associated hospitalization and severe disease also accounted for the presence of underlying risk factors.

**Figure S5.**
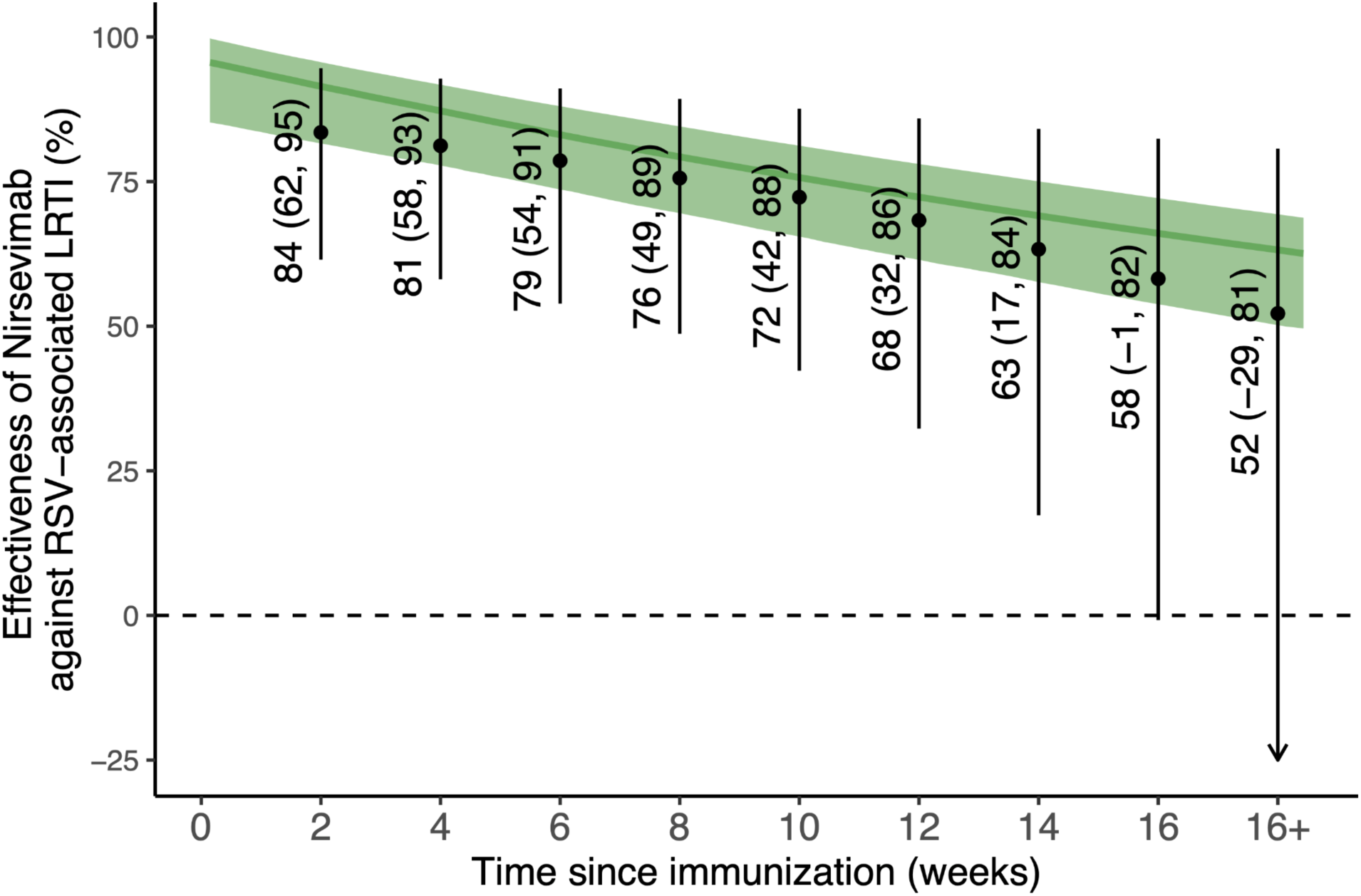
Effectiveness of nirsevimab against RSV-associated LRTI by time since immunization. The green curve and shaded area represent the median and 95% credible interval of the estimated efficacy of nirsevimab reported by Hodgson et al. [2], where efficacy over time was estimated using data from Phase IIb and Phase III trials in a survival model. The black dots denote the median estimates of nirsevimab’s effectiveness in preventing RSV-associated LRTI from our current study, using the same endpoint as in Hodgson et al. for comparison [2]. The error bars show the 95% credible intervals for the estimates, and the labels provide the exact values.

**Figure S6.**
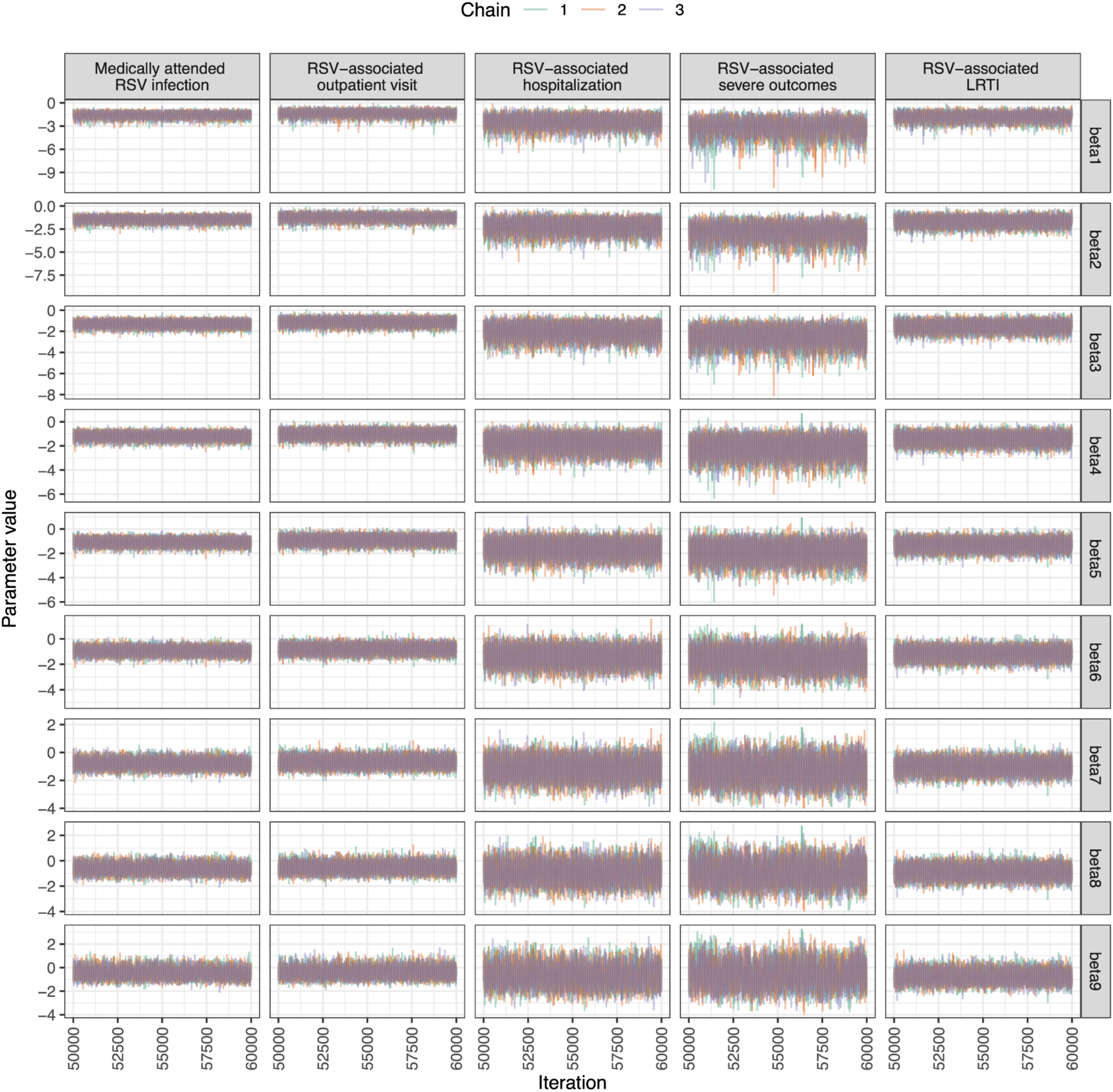
Trace plots for the coefficients of waning effectiveness. Trace plots for *β_n_* (effectiveness coefficient for each biweek interval after nirsevimab immunization, n = 1,2,3,…9) are displayed by the examined outcome. All parameters demonstrate good convergence. Iterations of the burn-in period (iterations 0–50,000) are excluded, and only the sampled iterations (50,000–60,000) are presented.

### SECTION 2. SUPPLEMENTAL TABLES REFERENCED IN THE MAIN MANUSCRIPT

**Table S1.** Definition of key clinical outcomes and risk factors.

| **Acute respiratory illness (ARI) [1]** | **Risk factors for severe RSV diseases** | **Medically attended upper respiratory infections (URTI)** | **Medically attended lower respiratory infections (LRTI)** |
| --- | --- | --- | --- |
| Acute onset (<10 days) illness that presents with at least two of the following: fever (measured or subjective), chills, rigors, myalgia, headache, sore throat, nausea or vomiting, diarrhea, fatigue, congestion, or one of the following: cough, shortness of breath, difficulty breathing, olfactory disorder, taste disorder, confusion, persistent chest pain, pale, gray, hypoxia, clinical or radiographic evidence of pneumonia or respiratory distress syndrome. | • Prematurity (gestational age less than 37 weeks)<br>• Anemia<br>• Immunodeficiency<br>• Cardiac abnormalities<br>• Pulmonary diseases<br>• Down syndrome<br>• Low birth weight (< 2,500 grams) | • Difficulty breathing<br>• Cough<br>• Croup<br>• Pain in throat / sore throat<br>• Nasal congestion<br>• Acute obstructive laryngitis<br>• Laryngeal stridor<br>• Pharyngitis<br>• Nasopharyngitis<br>• Otitis media<br>• Reactive airway diseases<br>• Upper respiratory tract infections recorded problem list with or without specifying pathogen | • Wheezing<br>• Bronchiolitis<br>• Bronchospasm<br>• Laryngotracheobronchitis<br>• Acute chest syndrome<br>• Pneumonia<br>• Hypoxia<br>• Hypoxemia<br>• Lower respiratory tract infections recorded in the problem list with or without specifying pathogen |

**Table S2.** Variable selection for the multivariable logistic regression models. The first row for each outcome presents the full model with all potential confounders. In each subsequent row, one variable is removed per step, with the final row showing the confounders included in the final model. Immunization status was included a priori in all models, and the final model was selected based on the lowest Akaike information criterion (AIC) score.

| Outcome | Confounders | AIC |
| --- | --- | --- |
| Medically attended RSV infection | age_tested + month_tested + race_ethnicity + atleastone_risk_factor + insurance_type | 2,853.0 |
|  | - race_ethnicity | 2,849.0 |
|  | - insurance_type | 2,846.8 |
|  | - atleastone_risk_factor | 2,845.1 |
|  | **(Final model: age_tested + month_tested)** |  |
| RSV-associated outpatient visit | age_tested + month_tested + race_ethnicity + atleastone_risk_factor + insurance_type | 2,242.0 |
|  | - race_ethnicity | 2,238.1 |
|  | - insurance_type | 2,236.3 |
|  | - atleastone_risk_factor | 2,235.6 |
|  | **(Final model: age_tested + month_tested)** |  |
| RSV-associated hospitalization | age_tested + month_tested + race_ethnicity + atleastone_risk_factor + insurance_type | 555.2 |
|  | - race_ethnicity | 552.1 |
|  | - insurance_type | 550.4 |
|  | **(Final model: age_tested + month_tested + atleastone_risk_factor)** |  |
| RSV-associated severe outcomes | age_tested + month_tested + race_ethnicity + atleastone_risk_factor + insurance_type | 425.4 |
|  | - race_ethnicity | 419.5 |
|  | - insurance_type | 419.0 |
|  | **(Final model: age_tested + month_tested + atleastone_risk_factor)** |  |
| RSV-associated LRTI | age_tested + month_tested + race_ethnicity + atleastone_risk_factor + insurance_type | 836.7 |
|  | - race_ethnicity | 830.7 |
|  | - insurance_type | 829.4 |
|  | **(Final model: age_tested + month_tested + atleastone_risk_factor)** |  |
| RSV-associated LRTI hospitalization | age_tested + month_tested + race_ethnicity + atleastone_risk_factor + insurance_type | 297.7 |
|  | - race_ethnicity | 293.9 |
|  | - insurance_type | 293.0 |
|  | **(Final model: age_tested + month_tested + atleastone_risk_factor)** |  |

**Table S3.** Comparison of immunized and unimmunized patients.

| Characteristic | Overall, N = 3,090<sup>1</sup> | Immunized, N = 330<sup>1</sup> | Unimmunized, N = 2,760<sup>1</sup> | Standardized Mean Difference<sup>1</sup> |
| --- | --- | --- | --- | --- |
| **Sex** |  |  |  | 0.03 |
| Female | 1,317 (42.6%) | 138 (41.8%) | 1,179 (42.7%) |  |
| Male | 1,772 (57.3%) | 192 (58.2%) | 1,580 (57.2%) |  |
| (Missing) | 1 (0.0%) | 0 (0.0%) | 1 (0.0%) |  |
| **Age at testing (months)** |  |  |  | -0.84 |
| Median (IQR) | 6.7 (3.6, 9.7) | 3.4 (1.8, 6.0) | 7.1 (4.1, 10.0) |  |
| **Race and ethnicity** |  |  |  | 0.32 |
| Hispanic | 1,328 (43.0%) | 138 (41.8%) | 1,190 (43.1%) |  |
| White non-Hispanic | 820 (26.5%) | 68 (20.6%) | 752 (27.2%) |  |
| Black non-Hispanic | 533 (17.2%) | 86 (26.1%) | 447 (16.2%) |  |
| Other non-Hispanic<sup>2</sup> | 161 (5.2%) | 24 (7.3%) | 137 (5.0%) |  |
| Unknown | 248 (8.0%) | 14 (4.2%) | 234 (8.5%) |  |
| **Birth weight** |  |  |  | -0.3 |
| Median (IQR) | 3,214.3 (2,824.9, 3,563.8) | 3,099.6 (2,575.6, 3,483.7) | 3,234.4 (2,875.0, 3,573.7) |  |
| (Missing) | 783 (25.3%) | 26 (7.9%) | 757 (27.4%) |  |
| **Gestational age < 37 weeks** | 418 (13.5%) | 90 (27.3%) | 328 (11.9%) | 0.63 |
| (Missing) | 757 (24.5%) | 22 (6.7%) | 735 (26.6%) |  |
| **Pulmonary diseases** | 156 (5.0%) | 19 (5.8%) | 137 (5.0%) | 0.04 |
| **Cardiac diseases** | 152 (4.9%) | 33 (10.0%) | 119 (4.3%) | 0.22 |
| **Anemia** | 94 (3.0%) | 11 (3.3%) | 83 (3.0%) | 0.02 |
| **Having at least one risk factor<sup>3</sup>** | 750 (24.3%) | 123 (37.3%) | 627 (22.7%) | 0.32 |
| **Insurance type** |  |  |  | 0.24 |
| Private | 983 (31.8%) | 78 (23.6%) | 905 (32.8%) |  |
| Public | 2,088 (67.6%) | 252 (76.4%) | 1,836 (66.5%) |  |
| Uninsured | 19 (0.6%) | 0 (0.0%) | 19 (0.7%) |  |

Data are presented as median (IQR) for continuous measures and n/total (%) for categorical measures.
<sup>1</sup>Standardized mean difference: the difference in means between case and control participants in units of the pooled SD. Covariates with an absolute standardized mean difference greater than 0.2 were considered to have important imbalances.
<sup>2</sup>Including Asian, Pacific Islander, Middle Eastern or Northern American, American Indian, or Native American by self-reporting.
<sup>3</sup>Have at least one of the following conditions recorded in the infant's medical history or diagnosis records: 1) Anemia; 2) Immunodeficiency (e.g. transplantation history, leukemia, etc.); 3) Cardiac diseases (including congenital heart diseases diagnosed at birth or any reporting of heart conditions); 4) Pulmonary diseases; 5) Down syndrome; 6) Small for gestational age (birth weight < 2,500 grams); 7) Prematurity (gestational age less than 37 weeks).

**Table S4.** Clinical characteristics of RSV-positive cases.

| Characteristic | Overall, N = 680<sup>1</sup> | Unimmunized, N = 659 | Immunized, N = 21<sup>1</sup> |
| --- | --- | --- | --- |
| **Hospital admission** | 166 (24.4%) | 161 (24.4%) | 5 (23.8%) |
| **Duration of hospitalization (days)** |  |  |  |
| Median (IQR) | 1.0 (1.0, 2.0) | 1.0 (1.0, 2.0) | 3.0 (2.0, 3.0) |
| N missing (% missing) | 514 (75.6%) | 498 (75.6%) | 16 (76.2%) |
| **ICU admission** | 23 (3.4%) | 22 (3.3%) | 1 (4.8%) |
| **Duration of ICU admission (days)** |  |  |  |
| Median (IQR) | 3.1 (1.8, 7.3) | 3.5 (1.8, 7.4) | 2.4 (2.4, 2.4) |
| N missing (% missing) | 657 (96.6%) | 637 (96.7%) | 20 (95.2%) |
| **Required high-flow oxygen support** | 145 (21.3%) | 141 (21.4%) | 4 (19.0%) |
| **Upper respiratory tract infection (URTI)<sup>2</sup>** | 229 (33.7%) | 221 (33.5%) | 8 (38.1%) |
| **Lower respiratory tract infection (LRTI)<sup>3</sup>** | 361 (53.1%) | 350 (53.1%) | 11 (52.4%) |
| **Fever (> 38°C/100.4°F)** | 239 (35.1%) | 235 (35.7%) | 4 (19.0%) |
| **Cough** | 98 (14.4%) | 94 (14.3%) | 4 (19.0%) |
| **Wheezing** | 9 (1.3%) | 9 (1.4%) | 0 (0.0%) |
| **Breathing difficulties** | 3 (0.4%) | 3 (0.5%) | 0 (0.0%) |
| **Bronchiolitis** | 349 (51.3%) | 338 (51.3%) | 11 (52.4%) |
| **Sepsis** | 2 (0.3%) | 2 (0.3%) | 0 (0.0%) |

<sup>1</sup>Data are presented as median (IQR) for continuous measures and n/total (%) for categorical measures.
<sup>2,3</sup> See Table S1 for definitions of URTI and LRTI.

**Table S5.**
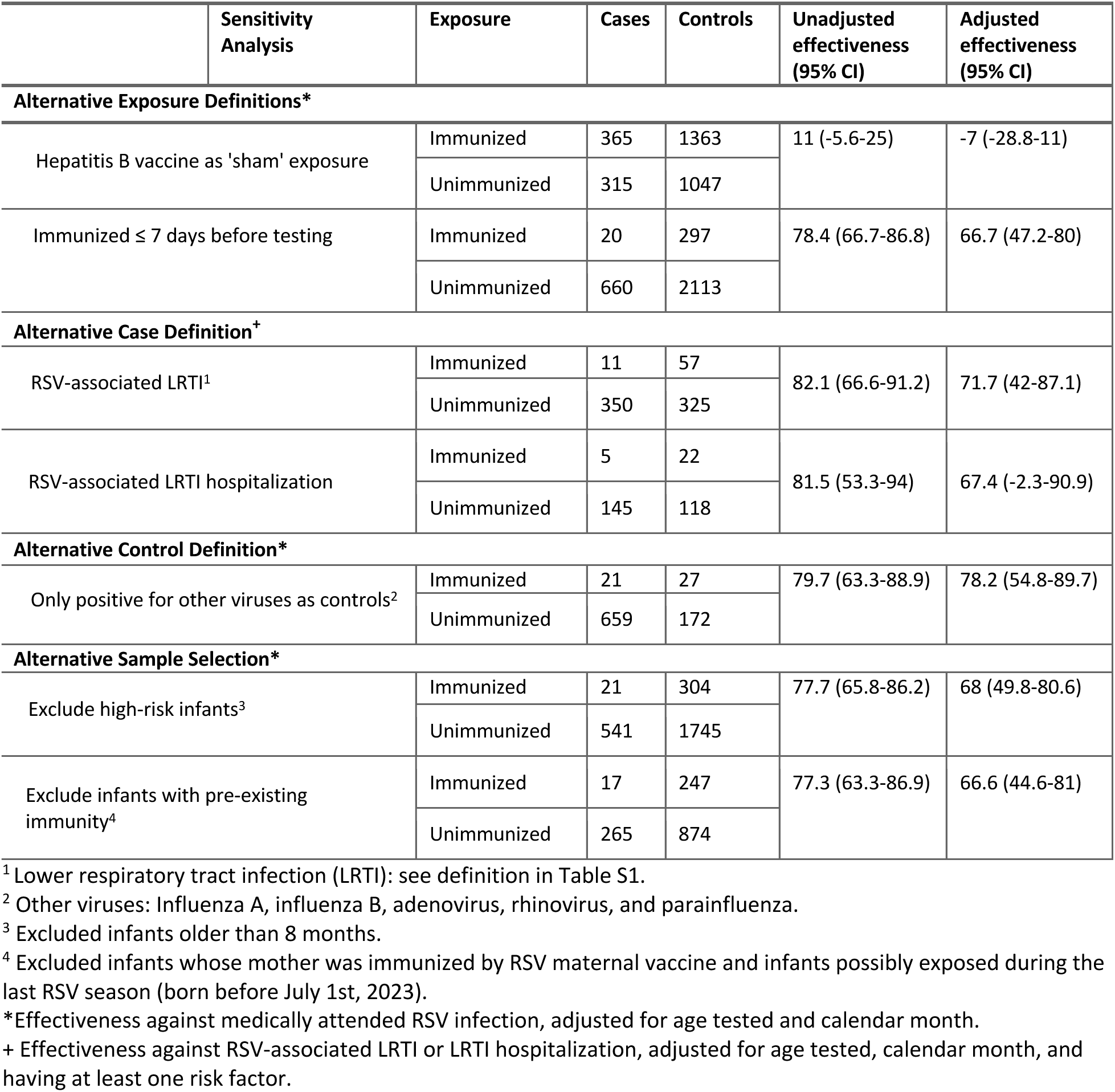
Sensitivity Analysis.

### SECTION 3. SUPPLEMENTAL METHODS

#### Estimating the effectiveness of nirsevimab over time

We evaluated the waning of nirsevimab’s protective effect over time using a logistic regression model within a Bayesian framework. For the jth test record in our dataset, the observed case status (i.e., whether the patient tested positive or negative for RSV) followed a Bernoulli distribution, such that

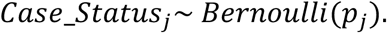

The time between vaccination with nirsevimab and sample collection for RSV testing was categorized into nine bi-weekly intervals (2, 4, 6, … >16 weeks). To incorporate this variable in the regression model, we created dummy variables to represent each time category (*time_since_vax_jn_, n* = 1,2,3 … 9). For example, if an individual was vaccinated 0-2 weeks before testing, *time_since_vax_j1_* = 1 and *time_since_vax_jn_* = 0 for *n* = 2,3, … 9. For an unvaccinated individual, all the dummy variables *time_since_vax_jn_* (*n* = 1,2,3, … 9) took the value of 0. The probability of testing positive, *p_j_*, was modeled using a multivariate logistic regression framework as follows:

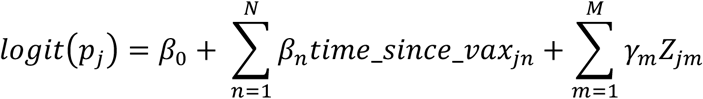

where *Z_jm_* represents potential confounders (e.g., age at testing, month of testing, risk factors), and *γ_m_* is the coefficients for the confounders.

Due to the waning nature of passive immunity, we assumed that nirsevimab’s effectiveness had a non-increasing trend over time. To reflect this in the model, we imposed a monotonic structure on the regression coefficients *β_n_*’s, such that

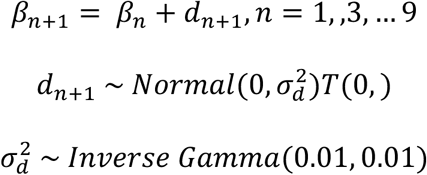

*T*(0, ) represents truncation at 0, allowing 𝑑_𝑛+1_ to take only non-negative values. For 𝛽_1_ (coefficient for the effectiveness 0-2 weeks after vaccination), we used a weakly informative prior distribution:

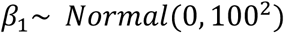

We examined effectiveness over time against various clinical endpoints. The model for each endpoint was fitted separately in the rjags package in R version 4.3.1, in which we collected 10,000 samples from the posterior distribution after discarding the first 50,000 samples in the burn-in period. Convergence was evaluated using trace plots (Figure S6). The estimated effectiveness of nirsevimab after a given period of time (for time interval n) since immunization *IE_n_* was calculated as

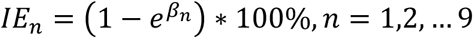

Medians and 95% quantile-based credible intervals were calculated from the collected posterior samples.

#### STROBE Statement—Checklist of items that should be included in reports of ***case-control studies***

|  | Item No | Recommendation | Page No |
| --- | --- | --- | --- |
| Title and abstract | 1 | (a) Indicate the study’s design with a commonly used term in the title or the abstract | 1,3 |
|  |  | (b) Provide in the abstract an informative and balanced summary of what was done and what was found | 3 |
| Introduction |  |  |  |
| Background/rationale | 2 | Explain the scientific background and rationale for the investigation being reported | 4 |
| Objectives | 3 | State specific objectives, including any prespecified hypotheses | 4 |
| Methods |  |  |  |
| Study design | 4 | Present key elements of study design early in the paper | 4 |
| Setting | 5 | Describe the setting, locations, and relevant dates, including periods of recruitment, exposure, follow-up, and data collection | 4-5 |
| Participants | 6 | (a) Give the eligibility criteria, and the sources and methods of case ascertainment and control selection. Give the rationale for the choice of cases and controls | 5 |
|  |  | (b) For matched studies, give matching criteria and the number of controls per case | NA |
| Variables | 7 | Clearly define all outcomes, exposures, predictors, potential confounders, and effect modifiers. Give diagnostic criteria, if applicable | 5 |
| Data sources/measurement | 8* | For each variable of interest, give sources of data and details of methods of assessment (measurement). Describe comparability of assessment methods if there is more than one group | 5 |
| Bias | 9 | Describe any efforts to address potential sources of bias | 5-6 |
| Study size | 10 | Explain how the study size was arrived at | 5 |
| Quantitative variables | 11 | Explain how quantitative variables were handled in the analyses. If applicable, describe which groupings were chosen and why | 5 |
| Statistical methods | 12 | (a) Describe all statistical methods, including those used to control for confounding | 5-6 |
|  |  | (b) Describe any methods used to examine subgroups and interactions | NA |
|  |  | (c) Explain how missing data were addressed | NA |
|  |  | (d) If applicable, explain how matching of cases and controls was addressed | NA |
|  |  | (e) Describe any sensitivity analyses | 6 |
| Results |  |  |  |
| Participants | 13* | (a) Report numbers of individuals at each stage of study—eg numbers potentially eligible, examined for eligibility, confirmed eligible, included in the study, completing follow-up, and analysed | 7 |
|  |  | (b) Give reasons for non-participation at each stage | NA |
|  |  | (c) Consider use of a flow diagram | 10 |
| Descriptive data | 14* | (a) Give characteristics of study participants (eg demographic, clinical, social) and information on exposures and potential confounders | 7, 14 |
|  |  | (b) Indicate number of participants with missing data for each variable of interest | 14 |
| Outcome data | 15* | Report numbers in each exposure category, or summary measures of exposure | 14 |
| Main results | 16 | (a) Give unadjusted estimates and, if applicable, confounder-adjusted estimates and their precision (eg, 95% confidence interval). Make clear which confounders were adjusted for and why they were included | 7,11 |
|  |  | (b) Report category boundaries when continuous variables were categorized | NA |
|  |  | (c) If relevant, consider translating estimates of relative risk into absolute risk for a meaningful time period | NA |
| Other analyses | 17 | Report other analyses done—eg analyses of subgroups and interactions, and sensitivity analyses | 8 |
| **Discussion** |  |  |  |
| Key results | 18 | Summarise key results with reference to study objectives | 8 |
| Limitations | 19 | Discuss limitations of the study, taking into account sources of potential bias or imprecision. Discuss both direction and magnitude of any potential bias | 9 |
| Interpretation | 20 | Give a cautious overall interpretation of results considering objectives, limitations, multiplicity of analyses, results from similar studies, and other relevant evidence | 9 |
| Generalisability | 21 | Discuss the generalisability (external validity) of the study results | 9 |
| **Other information** |  |  |  |
| Funding | 22 | Give the source of funding and the role of the funders for the present study and, if applicable, for the original study on which the present article is based | 1 |

\*Give information separately for cases and controls.
**Note:** An Explanation and Elaboration article discusses each checklist item and gives methodological background and published examples of transparent reporting. The STROBE checklist is best used in conjunction with this article (freely available on the Web sites of PLoS Medicine at http://www.plosmedicine.org/, Annals of Internal Medicine at http://www.annals.org/, and Epidemiology at http://www.epidem.com/). Information on the STROBE Initiative is available at http://www.strobe-statement.org.

